# Unsupervised Dimensionality Reduction Techniques for the Assessment of ASD Biomarkers

**DOI:** 10.1101/2024.08.12.24311682

**Authors:** Zachary Jacokes, Ian Adoremos, Arham Rameez Hussain, Benjamin T. Newman, Kevin A. Pelphrey, John Darrell Van Horn, the ACE GENDAAR Consortium

**Affiliations:** School of Data Science, University of Virginia; College of Computer, Mathematical, and Natural Sciences, University of Maryland; The Human Genetics Branch, National Institute of Mental Health; Department of Psychology, University of Virginia; Department of Neurology, University of Virginia Charlottesville, VA 22903, United States of America

**Keywords:** Autism, Neuroimaging, Copy number variation, Gene expression, Conduction velocity

## Abstract

Autism Spectrum Disorder (ASD) encompasses a range of developmental disabilities marked by differences in social functioning, cognition, and behavior. Both genetic and environmental factors are known to contribute to ASD, yet the exact etiological factors remain unclear. Developing integrative models to explore the effects of gene expression on behavioral and cognitive traits attributed to ASD can uncover environmental and genetic interactions. A notable aspect of ASD research is the sex-wise diagnostic disparity: males are diagnosed more frequently than females, which suggests potential sex-specific biological influences. Investigating neuronal microstructure, particularly axonal conduction velocity offers insights into the neural basis of ASD. Developing robust models that evaluate the vast multidimensional datasets generated from genetic and microstructural processing poses significant challenges. Traditional feature selection techniques have limitations; thus, this research aims to integrate principal component analysis (PCA) with supervised machine learning algorithms to navigate the complex data space. By leveraging various neuroimaging techniques and transcriptomics data analysis methods, this methodology seeks to contextualize the complex genetic and phenotypic heterogeneity linked to sex differences in ASD and pave the way for tailored interventions.

## 1. Introduction

Autism Spectrum Disorder (ASD) encompasses a broad range of developmental conditions characterized by deficits in social functioning, cognition, and behavior^1^. Individuals with ASD often experience challenges in communication, social interactions, and may engage in repetitive behaviors or have narrowly focused interests^2^. The prevalence of ASD has been steadily increasing worldwide, affecting between 1 in 36 children and 1 in 45 children according to recent meta-analyses and research by the Centers for Disease Control and Prevention (CDC)^3,4^. This diagnostic increase has brought significant attention to the urgent need for a deeper understanding of the underlying mechanisms of ASD.

Research indicates that ASD is a heterogeneous condition, meaning that it can present very differently from one person to another, complicating efforts to pinpoint its causes^5^. Although it is widely accepted that both genetic and environmental factors contribute to the development of ASD, the exact etiological factors and their interactions remain unclear. Genetic studies have identified numerous genes associated with ASD, suggesting a strong hereditary component^6–8^. However, environmental factors such as prenatal exposure to certain drugs, complications during birth, and advanced parental age have also been identified as potential risk factors for developing ASD^9,10^.

Moreover, neuroimaging studies have revealed differences in brain structure and function in individuals with ASD^11–13^. These studies have shown abnormalities in areas of the brain responsible for social behavior, communication, and sensory processing. Despite these advances, there is still much to learn about how these genetic and environmental factors interact to influence brain development and lead to the diverse array of symptoms observed in ASD.

Neuroimaging and genomics exploration is essential for understanding ASD because these approaches provide complementary insights into the biological underpinnings of the condition. Neuroimaging techniques, such as MRI and fMRI, allow researchers to observe structural and functional differences in the brains of individuals with ASD; this imaging data helps to identify patterns and variations in brain development and connectivity that may contribute to ASD symptoms. Concurrently, genomics offers a window into the genetic factors influencing ASD risk, uncovering specific genes and genetic variants associated with the disorder. By integrating genomic information with neuroimaging data, research efforts can better explore how genetic predispositions affect brain structure and function, and vice versa. This combined approach is crucial for elucidating the complex interplay between genetic and neural mechanisms, ultimately enhancing our understanding of ASD and guiding the development of more targeted interventions.

### 1.1. Sex-wise disparity in ASD

A significant aspect of ASD research is the observed sex-wise disparity in its prevalence. Males are diagnosed with ASD more frequently than females, with a ratio of approximately four-to-one^3^. This disparity suggests potential sex-specific biological factors that may influence the development of ASD. Several hypotheses have been proposed to explain this difference, including genetic differences in sex chromosomes, hormonal influences, and differences in brain structure and function between males and females^12,14^. Understanding these sex-specific factors is crucial for developing tailored diagnostic and therapeutic approaches for ASD.

### 1.2. Neuronal microstructure analysis in ASD

Neuroscientific research has increasingly focused on the neuronal microstructure to uncover the subtle differences in brain form and function associated with ASD. Using diffusion MRI, microstructural analysis allows for the examination of small-scale variations in the brain’s cellular architecture and can provide insights into the neural underpinnings of ASD. A recently developed microstructural analysis measures axonal conduction velocity, which is derived from parameters such as the g-ratio (the ratio of the inner to the outer diameter of the myelin sheath) and axon diameter^15,16^. Conduction velocity approximates the speed at which action potentials travel along axons, and deviations from the optimal speed can result in impaired neuronal communication.

### 1.3. Genetic factors and the pseudo-autosomal region

Genetic research has identified several candidate genes associated with ASD, many of which are in the pseudo-autosomal regions of the sex chromosomes^17–19^. These regions are of particular interest because they escape the usual X-inactivation process in females, resulting in a unique expression pattern that may contribute to the sex-wise disparity observed in ASD. Exploring these genetic factors, combined with microstructural data, can provide a more comprehensive understanding of the biological basis of ASD.

### 1.4. The ACE Network and NDA

The Autism Centers of Excellence (ACE) program is an initiative funded by the National Institute of Mental Health (NIMH) aimed at advancing the understanding, diagnosis, and treatments of ASD. Established to support large-scale multidisciplinary research projects, the ACE program brings together leading experts from various fields like genetics, neuroimaging, and phenotypic science to foster collaboration. Its structure allows for the integration of novel methodologies and state-of-the-art technologies to ensure that research efforts are at the forefront of scientific discovery. Complementary to the ACE program is the NIMH Data Archive (NDA), a comprehensive database managed by the NIMH that serves to centralize and disseminate the vast array of data collected on mental health research. Together, the ACE program and the NDA create a synergistic environment to nurture and advance the field of ASD research. The ACE program generates rich multimodal datasets that feed into the NDA. By leveraging the comprehensive data available through the NDA, researchers can explore new hypotheses, validate findings, and translate discoveries into clinical applications more effectively.

### 1.5. Dimensionality in microstructural analysis

A significant challenge in the analysis of neuronal microstructure data is the so-called “curse of dimensionality”. Microstructural processing pipelines typically generate data from over 200 distinct brain regions for each individual participant, which when performed on a voxel-wise level results in millions of datapoints for each individual. In our study, which includes 213 participants, this results in a vast multidimensional dataset. Although an N=213 might be considered respectable in human neuroimaging research, the sheer number of predictors poses a challenge for attaining sufficient statistical power, reproducibility, and interpretation. As an addendum to the concept of “big data,” we suggest that researchers consider highly dimensional datasets such as this one as “wide data” that is subject to a different set of equally important challenges.

Traditional approaches to address this issue involve feature selection to reduce the analytic search space. However, such techniques have inherent limitations. Firstly, they rely heavily on domain expertise, which may not always be available or infallible. Secondly, feature selection excludes certain predictors from the analysis before any machine learning algorithms can utilize them, thereby potentially limiting the scope of the analysis. While this approach can be beneficial when domain expertise is available, it can hinder exploratory analyses of new datasets.

### 1.6. Multimodal data fusion in health sciences

The integration of multimodal neuroimaging and genetic data presents a significant opportunity to improve model performance resulting from the synergy of shared and complementary information across modalities. For ASD research, the known genetic and neurological bases provide a strong foundation for exploring the rich multimodal data space afforded by large-scale data repositories like NDA provides for the ACE program. However, emphasizing interpretable methods is of paramount importance if research findings are to be translated into clinical application. It is through this framework this study has sought to provide insights into the multimodal data space generated by combining neuroimaging and genetic features.

### 1.7. Novel approach: PCA and machine learning integration

Our analysis aims to navigate the complex multidimensional space created by combining genetic and microstructural data modalities. To achieve this, we employ a novel implementation of principal component analysis (PCA) to identify unique characteristics of the dataset in an unsupervised manner. PCA allows us to reduce the dimensionality of the dataset while retaining the within-class variation, thereby addressing the curse of dimensionality without relying on traditional feature selection methods, as well as retaining generalizability to unseen data.

Following the unsupervised feature selection through PCA, we integrate the results into a traditional classification machine learning framework. This approach enables us to leverage the strengths of both unsupervised and supervised learning techniques, providing a more robust analysis of the data. By doing so, we aim to uncover novel insights into the relationship between genetic factors, neuronal microstructure, and ASD.

The integration of advanced neuroimaging techniques and genetic data analysis holds great promise for unraveling the complex etiology of ASD. By addressing the challenges posed by the curse of dimensionality and leveraging advanced analytical methods, we can enhance our understanding of how the neuronal microstructure and genetic factors combine to form the autistic phenotype. This comprehensive approach not only advances our knowledge of ASD but also paves the way for the development of more effective diagnostic and therapeutic strategies tailored to the unique needs of individuals with ASD.

## 2. Methods

### 2.1. Participants

Participants included 213 (mean age=153.20 [in months], standard dev.=±35.22; age range=96– 215; 99 female [46.48%]) volunteers from Wave 1 of an NIH-sponsored Autism Centers for Excellence network. The study sample included 113 autistic individuals (mean age=150.19, standard dev.=±34.56; age range=96–215; 51 female [45.13%]) and 100 non-autistic individuals (mean age=156.60, standard dev.=±35.81; age range=97–215; 48 female [48.00%]). The diagnostic and sex ratios were intended to be balanced. All ACE GENDAAR Wave 1 (9/04/2012-7/31/2022) neuroimaging, phenotypic, and genetic data were collected, processed, and archived on secure local compute servers under the following Internal Review Board (IRB) approvals: USC Approval #HS-13-00668; USC Approval #HS-18-00467; UVA Approval #22078; UVA IRB HSR #21361; GMU #00000169; and UVA #HSR-22-0423. As per the requirements of the US NIMH, de-identified and de-linked copies of all data were regularly submitted to the NDA as part of Collection #2021, where they are freely available for access to approved investigators. Data obtained by subsequent ACE GENDAAR Waves 2 and 3 (ongoing data collection) were not considered in this analysis. Informed consent was obtained from all participants and their legally authorized representatives.

### 2.2. Genetic data preparation

#### 2.2.1. Analysis of copy number variant densities

Using Bioconductor R, a karyotype map was created to visualize mutation densities^20^. Statistical differences were assessed between groups to determine mutation loci present in exclusively in ASD females, and vice versa. Loci were systematically compared to the locations of known genes using the UCSC genome browser, along with their exonic sections and prior association with ASD^21^. Copy number variants (CNVs) were identified from a set (N=196) of Manta-annotated variant-call format (VCF) files. The New York Genome Institute preprocessed and designed these files. Manta is a structural variant (SV) calling tool from Chen et al. that utilizes discordant read-pair and split-read evidence to identify various CNVs, including insertions, deletions, translocations, inversions, and tandem duplications^22^. Manta-annotated VCF files for each subject were compared against a Homo sapiens (assembly GRCh38.p14) reference genome, which contains base-pair positions for transcripts, genes, exons, and introns for all 24 chromosomes, including sex-linked chromosomes X and Y.

#### 2.2.2. Analysis of differential expression and functional enrichment analytics

Whole blood transcriptome sequencing was performed on 370 individuals. Transcript-level abundances were quantified using Kallisto^23^. Tximport was employed to aggregate these transcript-level abundances into gene-level counts^24^. Differential expression analysis was conducted using the R package DESeq2, facilitating the identification of statistically significant changes in gene expression across ASD-diagnosed individuals were compared across neurotypical cohorts with gender and diagnosis considered interaction effects^25^. Various functional enrichment analytics were conducted using multiple software packages and programs, including GSEA, clusterprofiler, Enrichr, and DAVID. Analytics such as gene-set enrichment analysis, gene ontology analysis, and KEGG pathway analysis were conducted on genes that are differentially expressed between individuals diagnosed with ASD and non-autistic cohorts.

### 2.3. Conduction velocity data preparation

#### 2.3.1. Image acquisition

Diffusion, T1-weighted, and T2-weighted images were acquired from each participant. Diffusion images were acquired with an isotropic voxel size of 2x2x2mm^3^, 64 non-colinear gradient directions at b=1000 s/mm^2^, and 1 b=0, TR=7300ms, TE=74ms. T1-weighted MPRAGE images with a FOV of 176x256x256 and an isotropic voxel size of 1x1x1mm^3^, TE=3.3; T2-weighted images were acquired with a FOV of 128x128x34 with a voxel size of 1.5x1.5x4mm^3^, TE=35. All images were preprocessed to correct for common sources of error and bias in accordance with prior published work^11,26^. T1w/T2w ratio was calculated by performing N4-bias correction, rescaling image intensity, then dividing on a voxel-wise basis^27,28^. Diffusion images were analyzed using a single-shell constrained spherical deconvolution (CSD) to obtain 3 tissue CSD (3T-CSD) microstructure compartments (intra- and extra-cellular isotropic signal, and intra-cellular anisotropic signal) and a fixel-based analysis was used to measure axonal fiber density and cross-section on a voxel-wise basis^11,26,29,30^. Despite obtaining multiple microstructure metrics using this methodology, only conduction velocity was examined here.

#### 2.3.2. Conduction velocity determination

The aggregate g-ratio was calculated on a voxel-wise basis and was used as Mohammadi & Callaghan suggest; this is displayed in Equation 1^16,31–33^. As a measure of intra-axonal volume, the fiber density cross section was used as the intra-axonal volume fraction (AVF), and as a metric of myelin density, the T1w/T2w ratio was used as the myelin volume fraction (MVF)^34^. Both metrics represent the total sums of each respective compartment across the volume of the voxel and are a volume-based equivalent to the original formulation of *g* as the ratio of axon diameter (*d*) to fiber diameter (*D*).

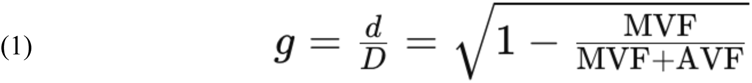

Aggregate conduction velocity was calculated based on the calculations of Rushton and Berman et al.; reiterating Rushton’s calculation that conduction velocity (*θ*) is proportional to the length of each fiber segment (*l*), and that this is roughly proportional to *D*, which in turn can be defined as the ratio between *d* and the g-ratio^15,35^. A value proportional to conduction velocity can be calculated using axon diameter and the g-ratio as in equation 2^35^:

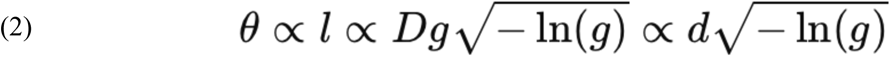

All imaging metrics, 3T-CSD compartments, T1w/T2w ratio, aggregate g-ratio, and aggregate conduction velocity were averaged across each of 214 ROIs taken from the JHU-ICBM WM atlas (48 ROIs) and the Destrieux Cortical Atlas (164 ROIs)^27,28^. Additionally, two composite ROIs were included, one of all 48 JHU ROIs and one of 150 neocortical regions from the Destrieux Atlas.

### 2.4. Initial analysis

#### 2.4.1. Data preprocessing

All conduction velocity and gene expression predictors were included in an initial traditional model for a total of 245 predictors. Participants were removed from the sample if missing either modality. The data was randomly split into training and testing sets, stratified by diagnostic cohort, at a 75-25 ratio. For feature preprocessing, all numeric predictors were normalized; the two modalities do not occur on the same scale, so in this way we ensured equitable contributions from each in the analysis.

#### 2.4.2. Principal component analysis

PCA was performed to reduce the dimensionality of the data. 40 principal components (PCs) were determined to be the maximum number of PCs examined: this number is equal to approximately 25% of the training data points (n=159), and 40 PCs account for approximately 85% of the cumulative explained variance.

#### 2.4.3. Logistic regression

Logistic regression modeling for classification was employed to determine how well the PCs separate the two classes. Model complexity was managed by tuning the number of PCs. 10-fold cross-validation was employed to further validate the modeling procedure. The workflow examined a range between one and 40 PCs to identify the number of PCs that maximized the area under the receiver operating characteristic (AUROC) curve, a metric that balances true positive rate against false positive rate. The final model configuration was applied to the entire training data set with the optimal hyperparameters determined by the tuning process. The final model was deployed on the unseen testing dataset, evaluated using both AUROC and accuracy. The results of the training and testing sets for this analysis are displayed in Table 1.

**Table 1.** Area under ROC curve and accuracy for the traditional model and experimental model.

|  | Training |  | Testing |  |
| --- | --- | --- | --- | --- |
|  | AUROC | Accuracy | AUROC | Accuracy |
| Traditional | 0.69306 | 61.26716% | 0.61813 | 57.40741% |
| Experimental | 0.69345 | 60.28922% | 0.66758 | 59.25926% |

### 2.5. Experimental analysis

#### 2.5.1. Data preprocessing

For the second comparative analysis, the existing training data set was split by participant cohort such that all autistic participants comprised one data frame and all non-autistic participants comprised another data frame. All conduction velocity and gene expression predictors were included in each of these data frames (again a total of 245 predictors). All numeric predictors were normalized again for the same reasons outlined above.

#### 2.5.2. Principal component analyses

Separate PCAs were performed on each of the cohort data frames to reduce the dimensionality of the cohort-specific data by exploring the underlying structures. The number of PCs retained were determined independently for each group. First, the number of PCs that account for 70% of the cumulative variance was identified. Then, the number of PCs with a corresponding eigenvalue greater than or equal to one was identified. If these numbers were not equal, the number of retained PCs was decided to be the midpoint between them (rounded down). The results of this process are displayed in scree plots in Figure 1. Consequently, 17 PCs were retained for the autistic cohort and 14 PCs were retained for the non-autistic cohort.

**Fig. 1.**
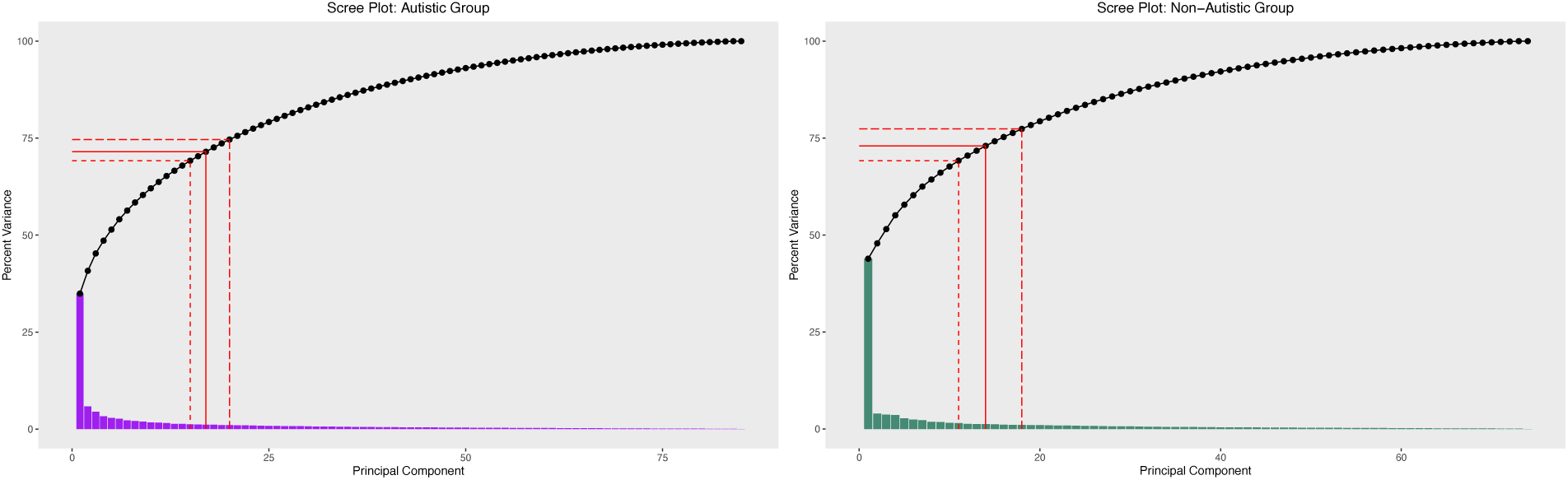
Scree plots of the autistic and non-autistic cohort PCAs. Short-dashed lines indicate the number of PCs that account for 70% of the cumulative variance, long-dashed lines indicate the number of PCs with eigenvalues greater than or equal to one; the solid red lines indicate the average of these two values, rounded down.

#### 2.5.3. Feature selection

Salient features for each group were extracted from the selected PCs systematically using the following procedure. First, the top 25% (75^th^ percentile) of variable loadings (in terms of absolute value) were identified per selected component to focus on those that contributed most to the within-class variance. Then, instances of each of the predictors present in the top 25% were aggregated to identify the unique predictors among and across these PCs, defined as those only appearing once across all selected PCs. This resulted in nine predictors for the autistic group and 31 for the non-autistic group. Finally, any common predictors between the two classes were removed and the remaining predictors were selected for modeling; this resulted in 36 predictors in total. A full accounting of these predictors is reported in Tables 2 and 3.

**Table 2.** Top predictors from the autistic cohort PCA, sorted by component number, then loading value within each component.

| Predictor | Region type | Hemisphere | Component | Value |
| --- | --- | --- | --- | --- |
| Frontal superior gyrus | Gray matter | Right | PC1 | 0.09903132 |
| Lateral fissure (posterior part) | Gray matter | Right | PC1 | 0.08902056 |
| Lateral superior temporal gyrus | Gray matter | Right | PC2 | 0.15112570 |
| Frontal inferior sulcus | Gray matter | Right | PC9 | -0.12629366 |
| Dorsal posterior cingulate gyrus | Gray matter | Left | PC9 | -0.10684352 |

**Table 3.** Top predictors from the non-autistic cohort PCA, sorted by component number, then loading value with each component.

| Predictor | Region Type | Hemisphere | Component | Value |
| --- | --- | --- | --- | --- |
| Superior corona radiata | White matter | Right | PC1 | 0.08975968 |
| Body of corpus callosum | White matter | - | PC1 | 0.08928453 |
| Posterior corona radiata | White matter | Right | PC1 | 0.08895067 |
| Anterior corona radiata | White matter | Left | PC1 | 0.08881578 |
| Posterior limb of internal capsule | White matter | Left | PC1 | 0.08863288 |
| Posterior thalamic radiation | White matter | Right | PC1 | 0.08812841 |
| Superior circular sulcus of the insula | Gray matter | Left | PC1 | 0.08767992 |
| Posterior corona radiata | White matter | Left | PC1 | 0.08751291 |
| Mid./posterior cingulate gyrus/sulcus | Gray matter | Right | PC1 | 0.08638030 |
| External capsule | White matter | Right | PC1 | 0.08592746 |
| Genu of corpus callosum | White matter | - | PC1 | 0.08496462 |
| Posterior thalamic radiation | White matter | Left | PC1 | 0.08466760 |
| Caudate | Subcortical | Left | PC1 | 0.08449230 |
| Sub-parietal sulcus | Gray matter | Left | PC1 | 0.08329393 |
| Precuneus gyrus | Gray matter | Left | PC1 | 0.08141795 |
| Superior temporal sulcus | Gray matter | Left | PC2 | -0.07436500 |
| Superior temporal gyrus (transverse) | Gray matter | Left | PC5 | -0.07551055 |
| Anterior circular sulcus of the insula | Gray matter | Right | PC5 | 0.06755295 |
| Inferior frontal sulcus | Gray matter | Left | PC7 | 0.10263944 |
| H-shaped orbital sulcus | Gray matter | Left | PC7 | 0.08055954 |
| Superior occipital gyrus | Gray matter | Right | PC8 | 0.08143941 |
| Hippocampus | Subcortical | Left | PC9 | -0.10926479 |
| Superior temporal gyrus (transverse) | Gray matter | Right | PC10 | 0.11044326 |
| Tapetum | White matter | Left | PC11 | -0.11043376 |
| Inferior parietal gyrus (supramarginal) | Gray matter | Left | PC12 | -0.13365818 |
| Paracentral lobule gyrus and sulcus | Gray matter | Right | PC13 | 0.15603998 |
| Transverse temporal sulcus | Gray matter | Left | PC14 | -0.10194675 |

#### 2.5.4. Logistic regression

Logistic regression modeling for classification was again employed to determine the effectiveness of this dimensionality reduction technique as compared to the traditional method. Predictors for this model included the 36 predictors selected from the procedure above. Model complexity was managed by tuning the number of PCs. 10-fold cross-validation was employed to further validate the modeling procedure. The workflow examined a range between one and 36 PCs to identify the number of PCs that maximized the AUROC curve. The final model configuration was applied to the entire training data set with the optimal hyperparameters determined by the tuning process. The final model was deployed on the unseen testing dataset, evaluated using both AUROC and accuracy. The results of the training and testing sets for this analysis are displayed in Table 1. All machine learning analyses and plot visualizations were created using the R package TidyModels^36^.

## 3. Results

### 3.1. Gene expression analysis

Differential expression analysis in DESeq2 showed that 3,707 genes exhibited significant differences in expression levels. Notably, several genes located near the pseudo-autosomal boundary and the heterochromatic regions of the Y chromosome displayed the most prominent discrimination between autistic and non-autistic participants. Among these, the homologously encoded zinc finger transcription factors ZFX and ZFY emerged as highly significant genes. After adjustment, ZFX and ZFY showed exceptionally low *p*-values.

### 3.2. Traditional analysis

The results of the traditional modeling procedure were as follows. The 10-fold cross validation procedure for tuning the number of principal components showed that the best training AUROC was 0.69306 at 12 PCs. The associated training accuracy was 61.26716%. For the unseen testing dataset, the AUROC was 0.61813, and the accuracy was 57.40741%. These values are reported in Table 1; ROC curves are displayed in Figure 2.

**Fig. 2.**
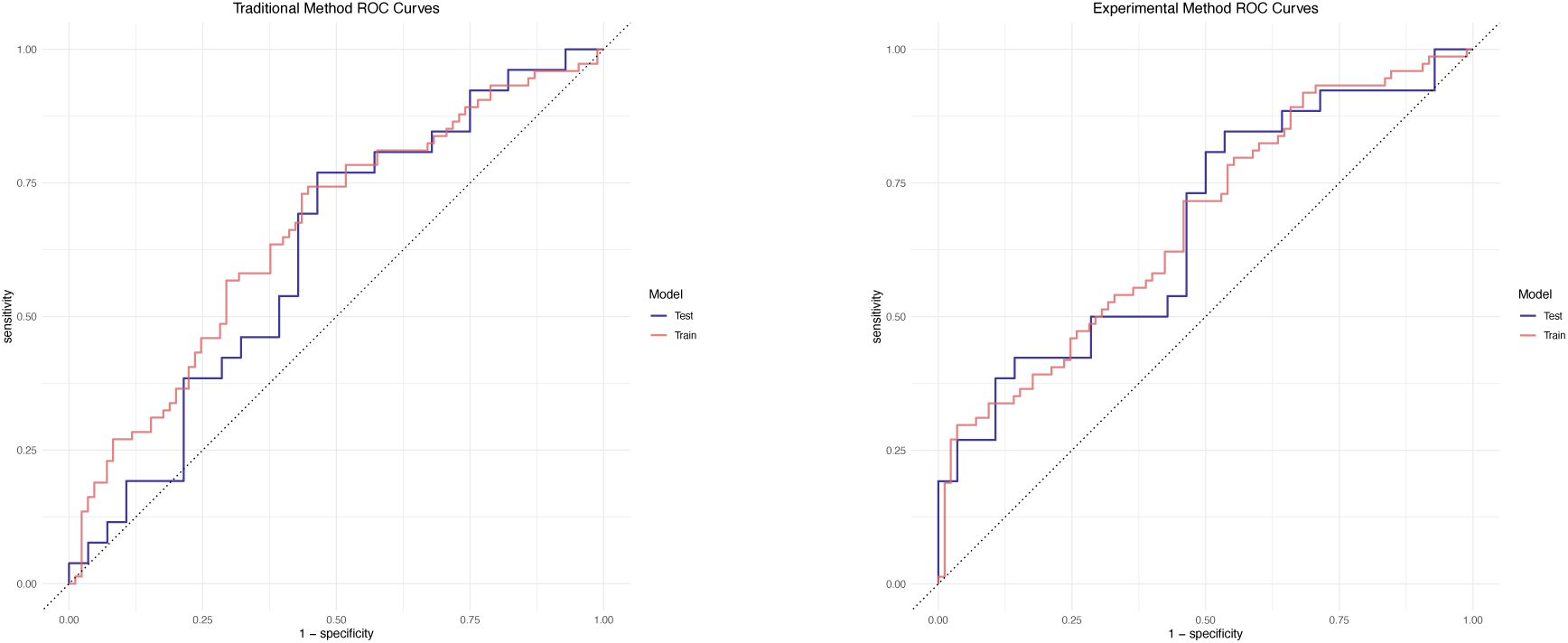
ROC curves for the traditional logistic regression results (left) and experimental results (right).

### 3.3. Experimental analysis

#### 3.3.1. Scree plot description

Scree plots were generated for each of the autistic and non-autistic cohort PCAs. Thresholds were determined based on the intersection of cumulative percent variance explained (greater than 70% were considered) as well as the principal components with eigenvalues greater than or equal to one; the average PC of these two metrics was used as the final threshold. These thresholds are shown in Figure 1; the former is indicated in smaller dashed red lines, the latter is indicated by longer dashed red lines, and the average of these two is also displayed as a solid red line. These values were as follows: greater than 70% cumulative variance was explained by 15 PCs in the autistic group and 11 PCs in the non-autistic group, eigenvalues greater than or equal to one included 20 PCs in the autistic group and 18 PCs in the non-autistic group, and the final threshold for the autistic group was 17 PCs and 14 PCs for the non-autistic group.

#### 3.3.2. Model evaluation

Table 1 contains the logistic regression performance results from the two approaches. The training AUROC and accuracy values were comparable across both approaches, while the testing AUROC of 0.66758 was greatly improved in the experimental approach, indicating more robust generalizability. Overall, the accuracy metrics were poor for both models, but an accuracy value of 59.25926% for the experimental approach showed improvement over the traditional approach. Visualizations of the AUROC curves are available in Figure 2.

#### 3.3.3. Feature selection

Tables 2 and 3 display the features selected by the experimental procedure, ordered by PC number and then loading value. After removing the predictors that appeared in PCA procedure for both the autistic and non-autistic group, the experimental analysis contained 36 predictors. These features were mostly loaded onto the first principal component for each group (25/40; 62.5%). The value reported in the final column of these tables represents the loading value of a given predictor on the PC where higher absolute values represent a stronger relationship between predictor and PC. Directionality is also relevant here: positive values indicate a positive relationship between predictor and PC, whereas negative values indicate the opposite. These values only apply within the context of a given PC and should not be compared across PCs. The relevant cortical, subcortical, and white matter regions can be found highlighted in Figure 3.

**Fig. 3.**
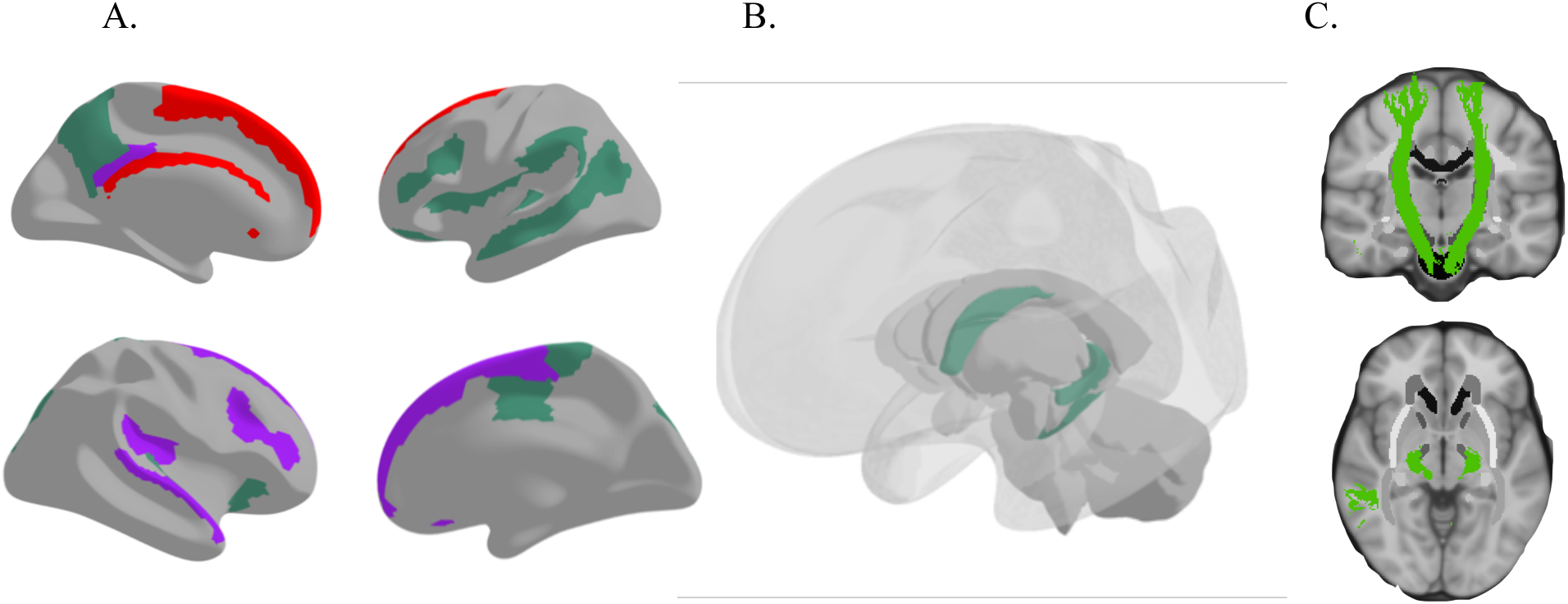
(A) Cortical regions extracted from the PCA procedure. Top left: medial view of the left hemisphere; top right: lateral view of the left hemisphere; bottom left: lateral view of the right hemisphere; bottom right: medial view of right hemisphere. (B) Subcortical regions extracted from the PCA procedure. (C) White matter tracts extracted from the PCA procedure. Purple regions were found to be characteristic of the autistic group; green regions were found to be characteristic of the non-autistic group; red regions represent the overlapping regions between both the autistic and non-autistic group.

## 4. Discussion

The results of the experimental dimensionality reduction procedure are promising. In the context of classification for autistic vs. non-autistic individuals using neuroimaging and genetic features, the AUROC performance in this study is acceptable, especially using traditional machine learning frameworks (and not deep learning, which brings its own set of challenges)^37–40^. PCA is effective in this context because it better addresses the issue of overfitting, as evidenced by the improved testing AUROC metric. By capturing within-class variability, the modeling effort performs better on unseen testing data and generalizes more readily to other datasets. Despite failing to achieve performance that could provide actionable clinical insights and true inference of the underlying mechanisms, the feature selection methodology succeeded for multiple reasons.

First, the marked improvement in testing AUROC performance (over the traditional approach) demonstrates that the extracted features capture many of the relevant aspects that differentiate the classes. AUROC is better suited to many classification tasks, including this one, since it provides a balance between true positive rate and false positive rate, whereas accuracy is a simpler metric that measures the ratio of correct predictions to total predictions. AUROC is also the preferred metric for datasets with imbalanced classes; while the classes in this study are not exceptionally imbalanced, AUROC is equipped to handle even slight imbalances and, as such, is the preferred metric here. AUROC improvements in the experimental analysis demonstrate this methodology’s internal validity and robustness to variations in unseen testing data.

Additionally, many of the extracted features represent notable regions of cortical, subcortical, and white matter connectivity in ASD research. ASD is characterized by abnormalities in brain structure, function, and connectivity, and many of the established areas of study are present in the extracted features^12,41,42^. The ability of the proposed procedure to pinpoint differences in relevant brain regions validates the methodology and necessitates further exploration both within and without the context of ASD research.

This analysis does not provide much evidence for the role of the pseudo-autosomal region on autism development, as none of the examined genetic predictors outperformed the microstructural predictors in terms of principal component loading. The low N of the sample is likely a contributing factor to this phenomenon, though it is also possible that the pseudo-autosomal region is not nearly as contributory to the etiology of ASD as microstructural metrics. Indeed, when the two modalities were examined separately, the genetic data performed poorly as predictors for classification within the same framework.

### 4.1. Feature selection

#### 4.1.1. Cortical features

Of the many cortical gray matter regions extracted by this methodology, two have been implicated in ASD research previously: the superior occipital gyrus and the frontal superior gyrus^43,44^. The frontal superior gyrus in particular is known to play a role in executive functioning, a domain previously identified as having deficits for autistic individuals relative to non-autistic individuals^45,46^. Other extracted cortical regions not directly implicated in ASD research do pertain to neurological processes relevant to areas previously identified as lacking in ASD individuals, including social cognition (anterior circular sulcus of the insula, inferior parietal gyrus), language processing (inferior frontal sulcus, superior temporal sulcus) and executive function (inferior frontal sulcus)^47–50^.

#### 4.1.2. Subcortical features

Subcortical features extracted using this method included the hippocampus and caudate nucleus. The hippocampus is known to be heavily involved in memory-related functions, and specific to ASD, both encoding and retrieval processes of episodic memory have been implicated as altered in ASD^51^. The caudate nucleus has been shown to have decreased connectivity in autistic individuals and is implicated in restricted and repetitive behavior development and increased autistic symptom severity as well^52–54^.

#### 4.1.3. White matter features

Many of the white matter features extracted in this study are also characteristic of the differences observed between autistic and non-autistic individuals. Corpus callosum tracts are most relevant here (body and genu of corpus callosum, superior/anterior/posterior corona radiata), but the tapetum has also been found to be under-connected in ASD relative to non-autistic individuals^55,56^.

### 4.2. Alternative approaches

#### 4.2.1. PCA procedure on different data frames

This experimental technique was deployed on this dataset in other ways to assess its effectiveness in different contexts. PCA was performed on each modality without first separating classes to attempt to capture modality-specific variability. Many of the extracted microstructure predictors remained the same as the focus of this study; however, this method also incorporated several genetic predictors as well. The resulting logistic regression yielded poor classification performance, likely due to an inability to extract the most salient features for each class.

Further, separate PCAs were performed on the four groups defined by the two different modalities and the two classes (autistic genetic, autistic microstructure, non-autistic genetic, non-autistic microstructure). Again, the microstructure metrics were comparable to those extracted in the main analysis of the study, and again this method allowed for more genetic predictors to contribute to the machine learning framework. This methodology performed even worse than before, however. The results of both attempts further cements the conclusion that the pseudo-autosomal region does not contribute to differences between autistic and non-autistic participants in this study and it is possible the genetic basis of ASD may lie elsewhere on the genome.

#### 4.2.2. Other machine learning models

Two other types of machine learning models were employed for the classification part of this analysis: random forest (RF) and quadratic discriminant analysis (QDA). These models are appropriate for data that is not expected to display a linear decision boundary and as such are more flexible. Logistic regression does expect the data to be linearly separable, and while that may appear to be a significant limitation of the modeling efforts of this study, RF and QDA performed far worse than logistic regression in both the traditional and experimental dimensionality reduction frameworks. One explanation for this could be that the data is not complete enough to allow for flexible models to generalize well. Microstructure and genetics are only two pieces of a larger puzzle that can include many other modalities like functional imaging, EEG, and behavioral data. Relatedly, while the extracted features comprise the major group differences in this dataset, they only capture part of the global within-group variability and therefore further limit the generalizability of the results; a phenomenon exacerbated by flexible machine learning methods.

### 4.3. Future directions

In the pursuit of assessing putative neurogenetic markers of ASD through the integration of neuroimaging, genomic, and phenotypic data, built upon the approach described here, several critical future directions emerge. One primary consideration is the utilization of data imputation to increase the sample size. While genetic data imputation may not be valid due to the potential introduction of biases and inaccuracies, it can be more appropriately applied to other metrics such as conduction velocity, pending further exploration and validation of the technique in this context. In terms of machine learning applications, while classification remains a viable approach, regression-based predictive modeling presents an avenue with the potential for more nuanced and informative results. Incorporating behavioral phenotyping outcome surveys, including measures of language, executive function, and social interaction, could provide rich data for these models, enhancing their predictive power and relevance.

An interesting observation from recent experimental models is the failure of the gene expression modality to contribute significantly following the feature selection procedure. When modalities were analyzed independently without considering diagnostic groups using principal component analysis for feature selection, the resulting classification performance was suboptimal compared to traditional methods. This issue was further compounded when PCA was applied separately to four interactive classes based on diagnostic groups and modalities (e.g., gene expression-autistic, gene expression-non-autistic). This suggests that the variance captured through the main PCA feature selection approach is sufficient for robust case classification, outperforming more granular feature selection strategies.

The feature selection approach could be applicable in individual nuances in autistic individuals; the initial provenance of salient features provides a starting point from which individual similarities and differences can be assessed. Additionally, sex-specific disparities in ASD are another critical area that warrants further examination and could be addressed by an exacting feature selection approach. Conducting separate PCAs for different sexes within the autistic group may reveal unique and actionable insights, potentially improving the performance of downstream machine learning models.

Moreover, several advanced analytical methods offer promising future directions:

- **Weighting predictors:** Assigning weights to predictors based on their loading values or the variance they explain on a logarithmic scale may enhance model performance.
- **Thresholding predictors:** Determining the number of predictors to retain from each principal component based on the distributional shape of loadings could improve feature selection.
- **Deep learning for data fusion:** Employing deep learning techniques to integrate multimodal data could capture complex relationships between neuroimaging, genomic, and phenotypic data. This is an emerging area with promising results but no unified optimal strategy yet^57,58^.

In summary, future research in the integration of neuroimaging, genomic, and phenotypic data in ASD will need to explore advanced data imputation techniques, leverage regression-based predictive modeling, and consider sex-specific analyses. Employing deep learning, sophisticated weighting and thresholding strategies, and advanced dimensionality reduction methods could significantly enhance the understanding and predictive power of these complex datasets.

### 4.4. Conclusions

The results of the experimental dimensionality reduction procedure for classifying autistic versus non-autistic individuals using neuroimaging and genetic features are promising. The AUROC performance achieved in this study is acceptable, especially within traditional machine learning frameworks. PCA effectively addresses overfitting, as indicated by the improved testing AUROC metric. By capturing within-class variability, the model performs better on unseen testing data and generalizes more readily to other datasets.

Firstly, the marked improvement in testing AUROC performance over traditional approaches indicates that the extracted features capture many relevant aspects differentiating the classes. AUROC is a balanced metric that accounts for both true positive and false positive rates, making it particularly suitable for datasets with even slight class imbalances. The improvements in AUROC demonstrate the methodology’s internal validity and robustness to variations in unseen testing data.

Many of the extracted features represent notable regions of cortical, subcortical, and white matter connectivity, which are well-documented in ASD research. Interestingly, the analysis did not provide substantial evidence for the role of the pseudo-autosomal region in autism development. None of the examined genetic predictors outperformed the microstructural predictors in terms of principal component loading. This may be due to the low sample size, but it also raises the possibility that the pseudo-autosomal region is not as contributory to the etiology of ASD as microstructural metrics. When examined separately, genetic data performed poorly as predictors for classification within the same framework, further supporting this conclusion.

Cortical features extracted from the analysis highlight critical regions involved in ASD, such as areas related to social cognition, language processing, and executive function. These regions are consistent with the existing literature on ASD, reinforcing their importance in understanding the disorder’s neurobiological underpinnings. Likewise, subcortical features identified include regions involved in emotion regulation, reward processing, and motor functions. Abnormalities in these areas are frequently reported in ASD studies, underscoring their relevance to the disorder’s phenotype and supporting the validity of the feature selection process. Finally, white matter features point to connectivity issues between different brain regions, which are a hallmark of ASD. Disruptions in white matter integrity can affect communication between cortical and subcortical regions, contributing to the diverse symptomatology of ASD.

Applying PCA to each modality without separating classes aimed to capture modality-specific variability. While some microstructure predictors remained consistent, this approach also included several genetic predictors. However, the resulting logistic regression yielded poorer than expected classification performance, likely due to an inability to extract the most salient features for each class. Separate PCAs for the four groups (autistic genetic, autistic microstructure, non-autistic genetic, non-autistic microstructure) also performed poorly, reaffirming that the pseudo-autosomal region may not significantly contribute to ASD classification.

Exploring more flexible machine learning methods, such as quadratic discriminant analysis and tree-based models, did not improve performance over logistic regression. This suggests that the proposed feature selection method is most effective with less flexible machine learning models, highlighting the need for careful selection of analytical techniques based on the data and research goals. The identification of critical cortical, subcortical, and white matter features aligns with existing ASD research, reinforcing their relevance in understanding the disorder’s neurobiological underpinnings. While the role of genetic predictors remains less clear, these findings highlight the need for meticulous selection of analytical techniques tailored to the specific characteristics of the data. Such comprehensive and data-driven strategies are vital for understanding the nuances of ASD and advancing for the field toward more effective and personalized diagnostics and interventions.

## Data Availability

Data used in this study is publicly available at: https://nda.nih.gov/edit_collection.html?id=2021

## Notes

### Competing Interest Statement

The authors have declared no competing interest.

### Funding Statement

This work was performed on behalf of the Autism Centers of Excellence GENDAAR Consortium (NIH R01 MH100028; Kevin Archer Pelphrey, Ph.D., PI).

### Author Declarations

All ACE GENDAAR Wave 1 (9/04/2012-7/31/2022) neuroimaging, phenotypic, and genetic data were collected, processed, and archived on secure local compute servers under the following Internal Review Board (IRB) approvals: USC Approval #HS-13-00668; USC Approval #HS-18-00467; UVA Approval #22078; UVA IRB HSR #21361; GMU #00000169; and UVA #HSR-22-0423. As per the requirements of the US NIMH, de-identified and de-linked copies of all data were regularly submitted to the NDA as part of Collection #2021, where they are freely available for access to approved investigators. Data obtained by subsequent ACE GENDAAR Waves 2 and 3 (ongoing data collection) were not considered in this analysis.

